# Predicting Onset of COVID-19 with Mobility-Augmented SEIR Model

**DOI:** 10.1101/2020.07.27.20159996

**Authors:** Neo Wu, Xue Ben, Bradley Green, Kathryn Rough, Srinivasan Venkatramanan, Madhav Marathe, Paul Eastham, Adam Sadilek, Shawn O’Banion

**Author notes:** Corresponding authors: {, }. equal contribution (Neo Wu, Xue Ben).

## Abstract

Timely interventions and early preparedness of healthcare resources are crucial measures to tackle the COVID-19 disease. To aid these efforts, we developed the Mobility-Augmented SEIR model (MA-SEIR) that leverages Google’s aggregate and anonymized mobility data to augment classic compartmental models. We show in a retrospective analysis how this method can be applied at an early stage in the COVID-19 epidemic to forecast its subsequent spread and onset in different geographic regions, with minimal parameterization of the model. This provides insight into the role of near real-time aggregate mobility data in disease spread modeling by quantifying substantial changes in how populations move both locally and globally. These changes would be otherwise very hard to capture using less timely data.

## Introduction

Following its initial spread in China, COVID-19 has quickly swept across many countries and developed into a global pandemic. As of July 14, 2020, there have been 12,964,809 confirmed cases globally, including 570,288 deaths [1]. This crisis has also brought serious challenges to many healthcare systems. Since the outbreak, many countries have been suffering from a shortage of healthcare professionals, personal protective equipment and other medical resources. Early warnings of a potential pandemic, including understanding where the disease will spread next, may inform resource allocation and early interventions by governments and public health organizations aimed at containment.

Human mobility data has played an important role in infectious disease modeling. Classic models such as the gravity model [2] and radiation model have been developed to model human movement in disease spreading. Colizza *et al*. developed metapopulation models based on reaction-diffusion theories characterized by heterogeneous connectivity and mobility patterns [4]. Balcan *et al*. integrated short- and long-term mobility flows to model spatiotemporal patterns of a global epidemic [5, 6]. Mobility-based models have been successfully applied to diseases such as influenza [5], malaria [7,8], measles [9], HIV [10] and SARS [11]. Similarly, human mobility data has already played a critical role in modeling COVID-19. For instance, Kraemer *et al*. analyzed the effect of human mobility and control measures on the COVID-19 epidemic in China [12]. Mobility data derived from air, road and rail traffic as well as social media were used to model the early outbreak of COVID-19 in China [13, 14] and early global spread [15]. More recently, Pei *et al*. developed a metapopulation model at county level in the United States and that employs two types of mobility flows, daily work commuting and random movement, to model the spread of COVID-19 [16].

In this paper, we present the Mobility-Augmented SEIR (MA-SEIR) model, an approach that is based on a variant of compartmental models [17, 18] and takes into account aggregate mobility data to predict COVID-19 spread across countries and states. In contrast to mobility data based on traffic networks or social media, Google’s aggregate and anonymized mobility data is derived from Google users who have turned on the Location History setting, which is off by default [19]. This is similar to the data used to show how busy certain types of places are in Google Maps — helping identify when a local business tends to be the most crowded. With this up-to-date mobility dataset, we show how the MA-SEIR model is able to forecast the onset of the COVID-19 epidemic at both global and local geographical levels, where the onset is defined as the time when the number of cases is above 100 at country level and 30 at US state level. We also illustrate its use to estimate the effect of reduced or increased mobility on COVID-19 spread by simulating different mobility scenarios. In the following sections we present retrospective analyses to test these claims.

## Results

### MA-SEIR predicts the onset of the global pandemic

The MA-SEIR parameters are estimated from the reported cases in China [14] (details in Supplementary Information). Since we aim to model how a novel infectious disease spreads across the globe, we assume good knowledge on parameterization is not available at the early stage of an epidemic. Nevertheless, we assume that a simple MA-SEIR model with time-*independent* parameters can capture the *early* stage of the epidemic reasonably well. When augmented with near real-time mobility data, we hypothesize that the MA-SEIR model can simulate the spread of COVID-19 and predict the onset of the epidemic in new geographic areas with minimal initial seeding and parameterization.

We assumed that COVID-19 outbreak started in Wuhan, China, thus the disease spread is simulated by seeding positive cases in China [20]. We experimented with a few seeding strategies, including seeding China with the earliest confirmed case, seeding cases in China and South Korea, and in China and Italy. We found that using the earliest confirmed case to seed the simulations yielded the best results in terms of interpretability and forecasting accuracy. This is likely due to the fact that different regions had inconsistent testing capacities early in the epidemic, and therefore seeding with confirmed cases from different regions may not accurately represent the earliest stage of the pandemic. Based on this analysis, we seeded all global simulations of country-level COVID-19 spread with only the first confirmed case in China on December 1, 2019, according to [20]. The simulations were run from December 1 to May 1, 2020, with real-time mobility flows up to January 30, 2020.

Figure 2 shows the performance of MA-SEIR in predicting the epidemic onset for a number of countries across the world. The countries shown in the figure are from six continents, including Asia, Europe, Africa, Oceania, North America, and South America. Countries below the dotted line show early predictions compared to the actual confirmed onset dates. We also note that, due to varying COVID-19 testing and reporting situations across the countries, the reported onset dates may be inaccurate with delays. Therefore, our prediction offers an alternative estimate of the actual epidemic across the world.

**Figure 1:**
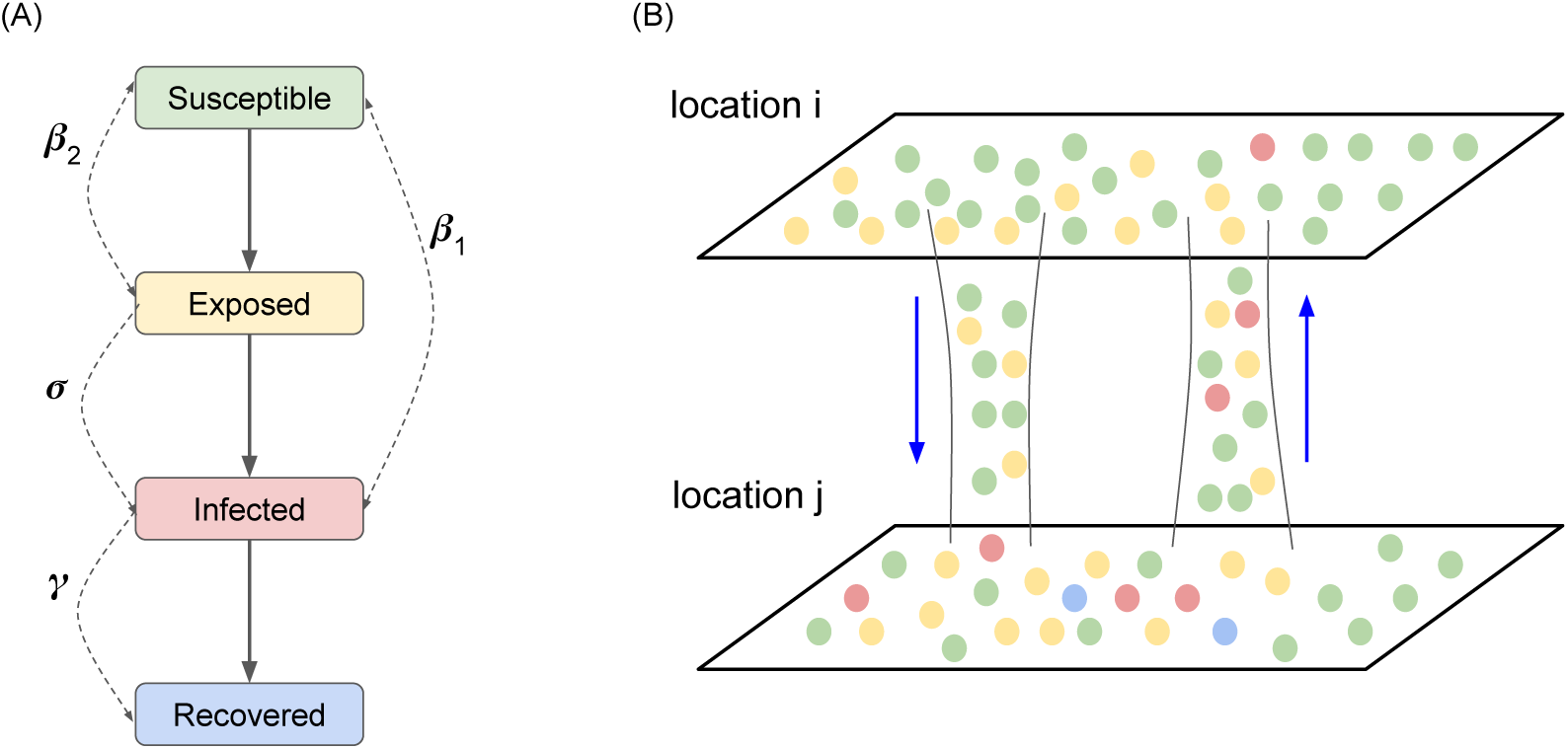
Illustration of the Mobility-Augmented SEIR (MA-SEIR) model.(A) The SEIR component of the MA-SEIR model. Each population is divided into four compartments, the Susceptible, Exposed, Infected and Recovered. The solid arrows indicate the allowed flows between the four states. The single- and double-headed dashed arrows denote the first- and second-order rate parameters of different interactions. (B) Each location is modeled as a metapopulation, whose dynamics is governed by the SEIR scheme. Inter-regional mobility data is used to model the interactions between two locations.

**Figure 2:**
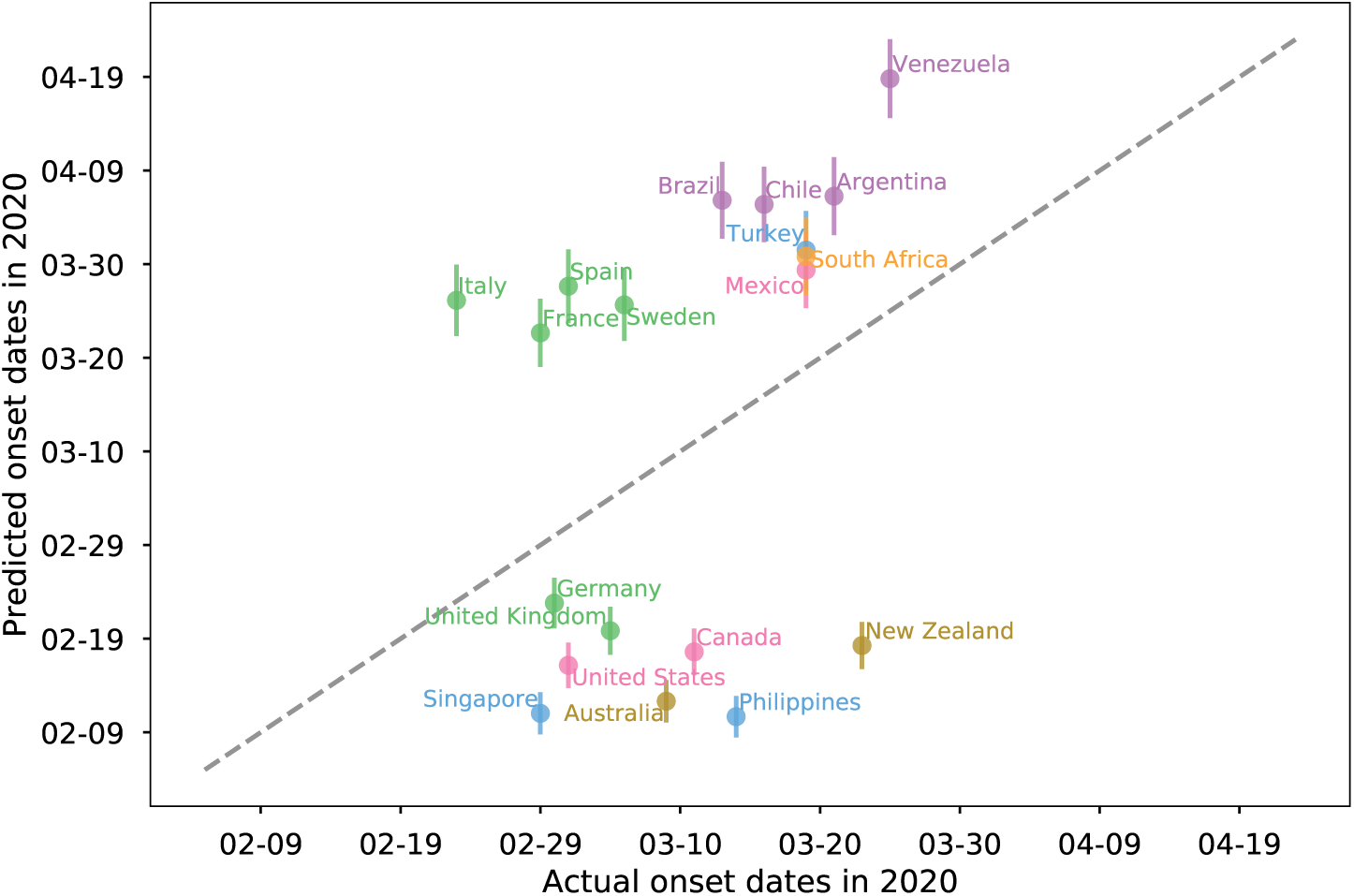
Predicted onset dates vs the actual onset dates for various countries across the world. The simulation is seeded with one case in China on December 1, 2019. The dots with the same color are from the same continent, including countries from Asia, Europe, Africa, Oceania, North America, and South America. The dotted line represents perfect matching predictions, where predicted days match exactly the actual onset dates. Error bars are included for the predicted onset dates.

Next we analyzed the forecasting performance of MA-SEIR for the 130 countries included in this study. Figure 4 shows the distributions of the errors in actual and predicted onset dates for 114 countries around the world. These are the countries that have seen epidemic onsets at the time of April 12, 2020 according to reported confirmed cases. Indeed the histogram suggests that the MA-SEIR model seeded with one case from December 1, 2019 is capable of predicting the epidemic onset in advance for most countries, as shown from the tail on the right side of the histogram. The summary metrics of the model performance on predicting the onset days are presented in Table 1. For countries with early onset predictions and late onset predictions, the corresponding mean absolute errors (MAEs) are 21.8 days and 18.8 days, respectively. Pearson correlation coefficient between the predicted and actual onset days is 0.51. Spearman’s rank correlation between the predicted and actual onsets is 0.67, suggesting a good positive correlation in predicting country onset in a relative manner.

**Table 1:** Metrics for country and state level predictions of COVID-19 onset.

| **Metrics** | **Median of Abs Error**<br>**(day)** | **Mean of Abs Error**<br>**(day)** |
| --- | --- | --- |
| Early predictions (global) | 21.2 | 21.8 |
| Late predictions (global) | 18.8 | 18.8 |
| All predictions (global) | 19.1 | 19.9 |
| Early predictions (US states) | 13.4 | 14.4 |
| Late predictions (US states) | 11.5 | 12.1 |
| All predictions (US states) | 11.9 | 12.2 |

**Figure 3:**
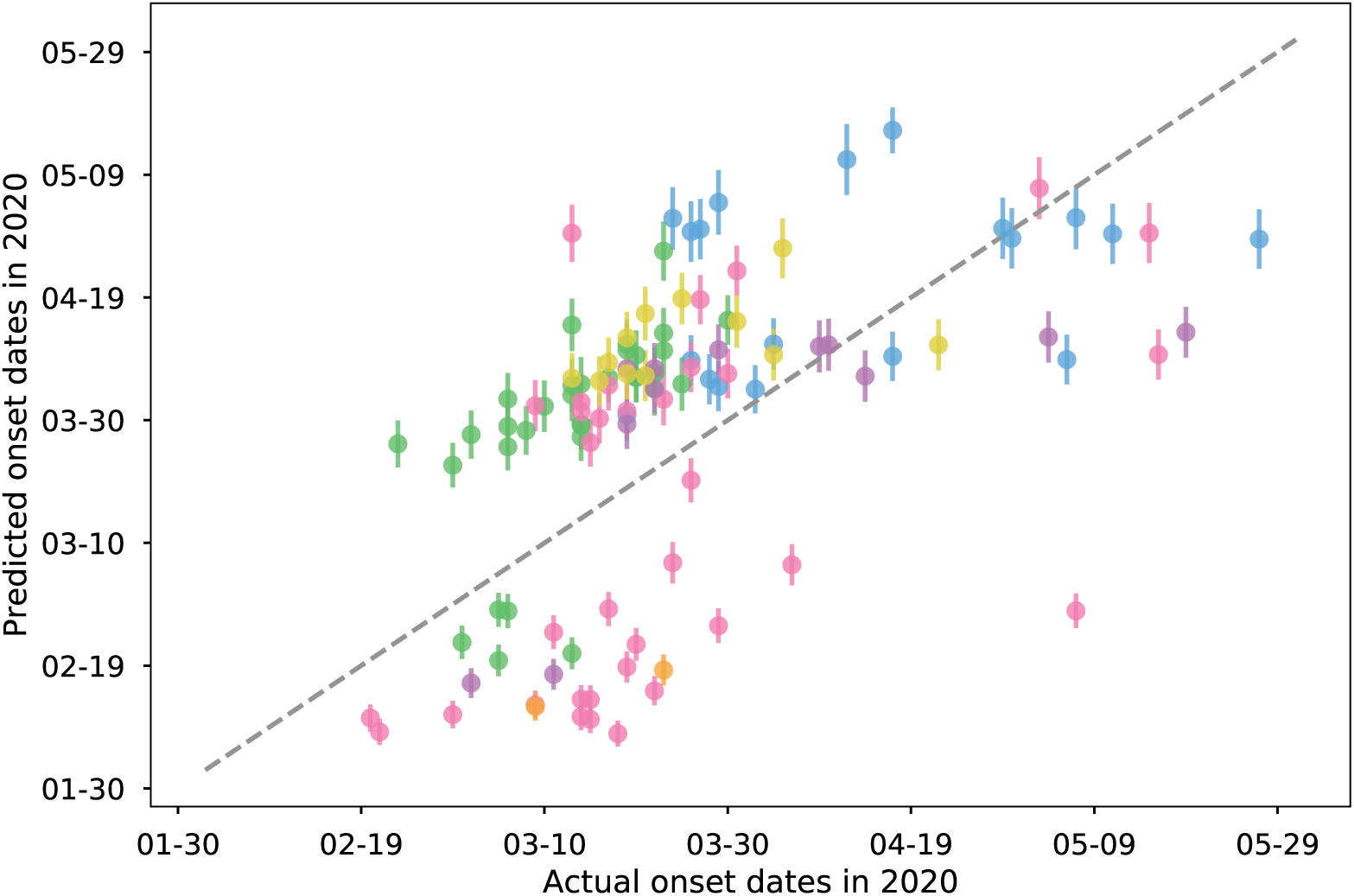
Predicted onset dates vs the actual onset dates for all countries across the world that have seen epidemic onset. The simulation is seeded with one case in China on December 1, 2019. The dots with the same color are from the same continent, including countries from Asia, Europe, Africa, Oceania, North America, and South America. Blue is Europe, green is Africa, pink is Asia, purple is North America, yellow is Oceania, orange is South America. The dotted line represents perfect matching predictions, where predicted days match exactly the actual onset dates. Pearson and Spearman correlation coefficient between the predicted and actual onset days is 0.51 and 0.67.

**Figure 4:**
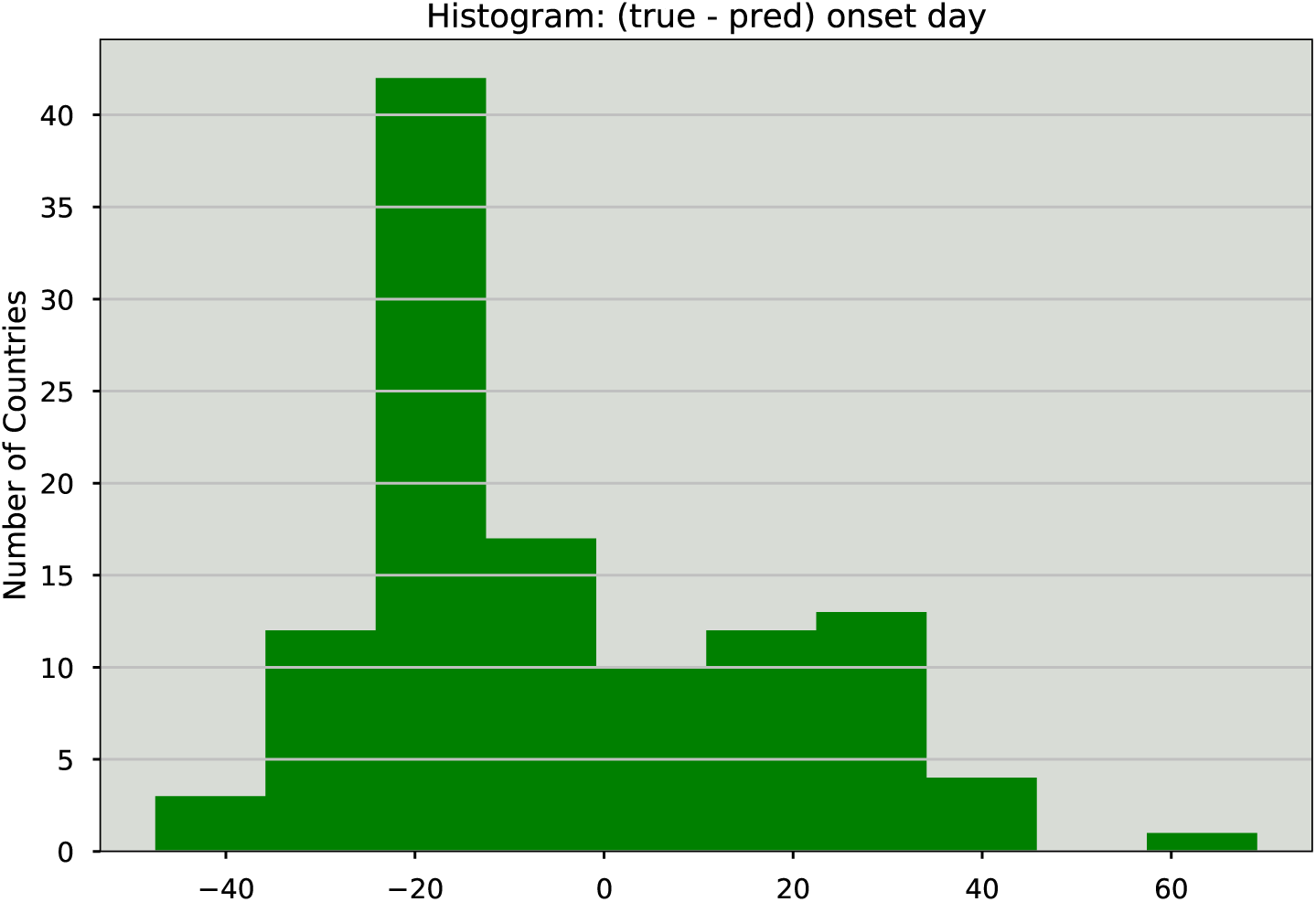
The distribution of errors between the true and predicted onset day for all countries that have seen epidemic onsets (total 114 countries), according to the reported confirmed cases. The simulation is seeded with one case in China on December 1st 2019. Positive error means early prediction from the model. Negative error means late prediction.

### MA-SEIR predicts regional epidemic outbreaks

We then tested the hypothesis whether MA-SEIR can forecast the onset of COVID-19 in each US state in a retrospective analysis. We seeded the simulation using 13 imported cases prior to February 14, 2020 (Table S1), and modeled the epidemics with aggregate and anonymized inter-state mobility flows. The simulations were run from January 23 to May 1, 2020, with real-time mobility flows up to February 14, 2020.

Figure 5 compares predicted and actual onset dates of the COVID-19 epidemic in each US state. The Pearson and Spearman correlation coefficients between the predicted onset and the actual date are 0.61 and 0.44, respectively. Overall, the MA-SEIR model predicted the onset of COVID-19 epidemic with good accuracy (MAE: 11.9 days). Among the 50 states, 35 states (shown in green in Figure 5) exhibit less than two weeks of delay in predicted onset time (MAE: 9.31 days). Thirteen states (Alabama, Georgia, Louisiana, Maryland, Mississippi, Nebraska, New Jersey, New York, North Dakota, Pennsylvania, South Carolina, South Dakota and Wyoming) exhibit delays between 2-3 weeks (shown in orange in Figure 5). Two states, Delaware and West Virginia, show significantly delayed onset (28 days for Delaware and 24 days for West Virginia). We next divided all the states into two groups based on whether the predicted onset is earlier or later than the reported onset, and computed the same metrics for each group. The MAEs are 13.4 days for the early prediction group and 11.5 days for the late prediction group. These results suggest that the MA-SEIR model is capable of forecasting the onset of local COVID-19 outbreak with reasonable accuracy, even though in some states the predicted onset date is delayed (Table S2).

**Figure 5:**
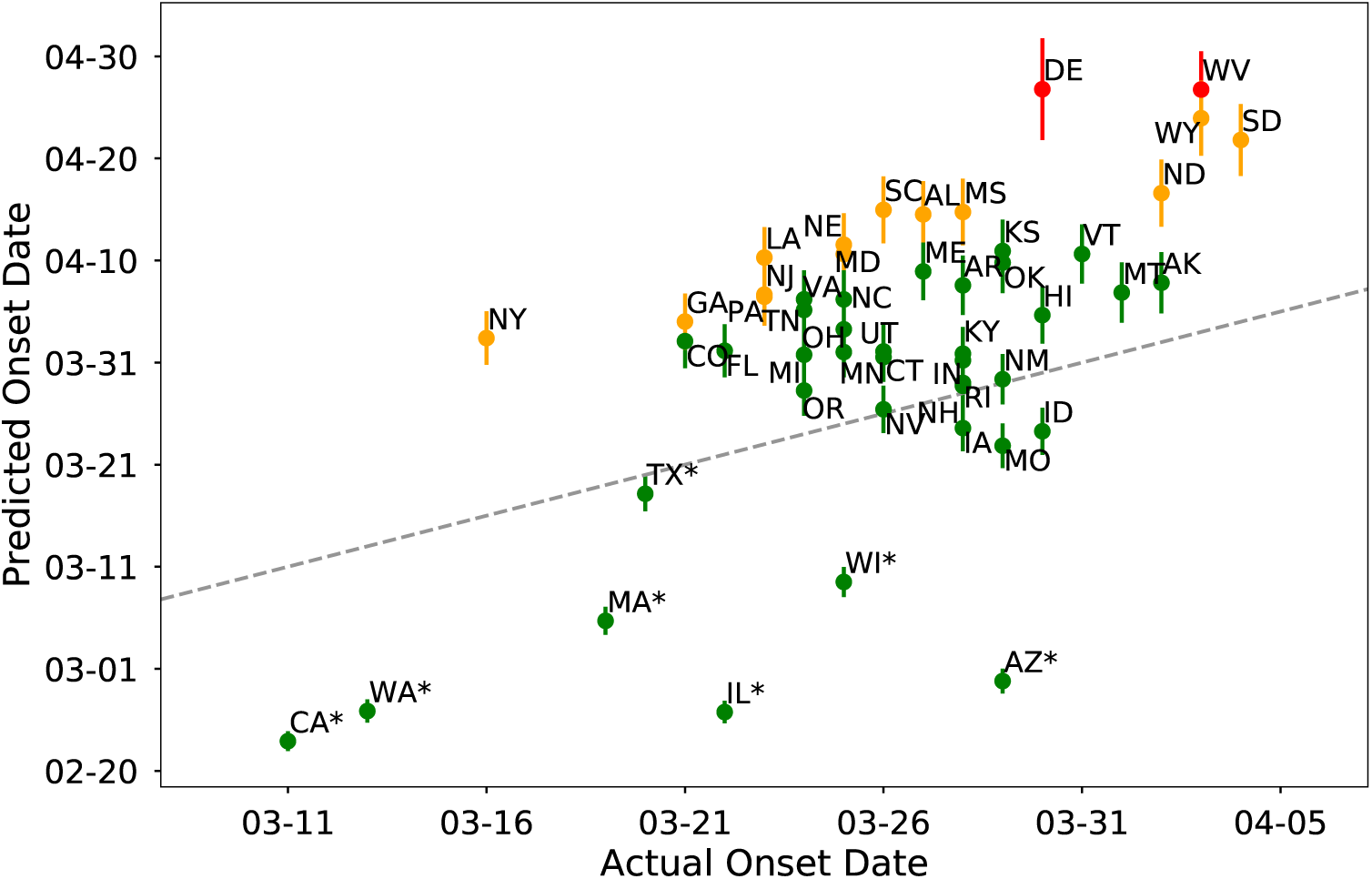
Comparison of predicted and actual onset dates for US states. The simulation is seeded with 13 confirmed cases between January 21, 2020 and February 14, 2020 in the United States. The states are colored by the delay in predicted onset compared to actual onset of the epidemic. Of all the states, 35 have less than two weeks of delay (green), 13 states show delays between 2-3 weeks (orange), and two states, Delaware and West Virginia, have more than three weeks of delay in predicted onset. Pearson correlation coefficient between predicted and actual onset dates is 0.61. The dotted line represents perfect matching predictions.

### MA-SEIR provides insights into mobility flows and disease spread

Here, we aimed to understand the relationship between the change of mobility and disease spread. We first analyzed the change of global mobility in response to COVID-19 outbreak. Starting from January 23, 2020, China has imposed strict lockdown measures to prevent the COVID-19 spread. Since then, many countries have adopted travel restrictions. As illustrated in Figure 6, the mobility flows in March 2020 have decreased substantially compared to that in January 2019 for the selected countries, especially countries in Asia and Europe. Italy experienced the strongest reduction among European countries. On average, the March mobility flows have dropped by 45.1% w.r.t. January 2020 and April mobility flows have dropped by 82.1%. These analyses suggest that travel restrictions have strongly suppressed the inter-country mobility.

**Figure 6:**
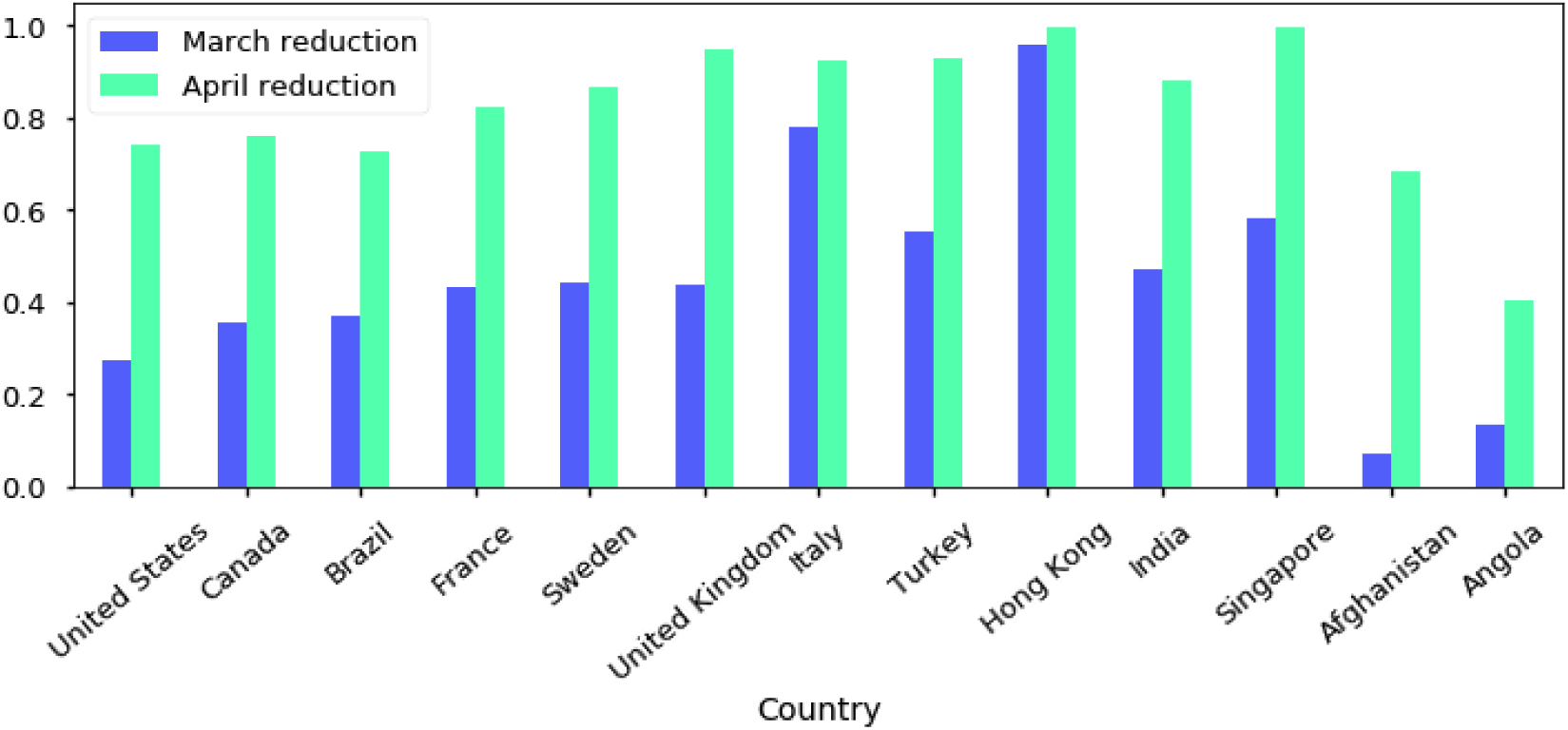
The relative reduction of inter-country mobility flows in Mar and April 2020 as compared to January 2020 for the selected countries. In March, countries in Asia and Europe have undergone substantial mobility reductions due to strict travel restrictions. The average reduction is 45.1% in March. In April, most countries have experienced substantial reductions with an average value of 82.1%.

To further understand how the change of inter-country mobility affects COVID-19 epidemic onset, we simulated the COVID-19 spread with reduced mobility flows. The simulated reduction occurred on December 21, 2019, that is, 20 days after the first seed in China.

We compared the predicted epidemic onset time using regular mobility flow versus using various levels of reductions in the flow, to evaluate the effects. We found that the mobility reduction indeed delayed the predicted epidemic onset time, as shown in Figure 7. The stronger the mobility reduction, the more the onset days were delayed. For countries, such as Spain, United Kingdom, Australia, Singapore, etc., the delay can be more significant under 85% mobility reductions. For Argentina, Brazil, etc., the delays don’t change with varying mobility reductions. We also selected the 130 countries that have seen epidemic onsets according to reported confirmed cases, and computed the average delays in the predicted onset days with different levels of mobility reductions. These analyses suggest that the effects of mobility reductions can be modeled by MA-SEIR and global mobility flows. While reduction of inter-country mobility can delay the onset of an epidemic, these effects are only significant with substantial reductions and vary by country.

**Figure 7:**
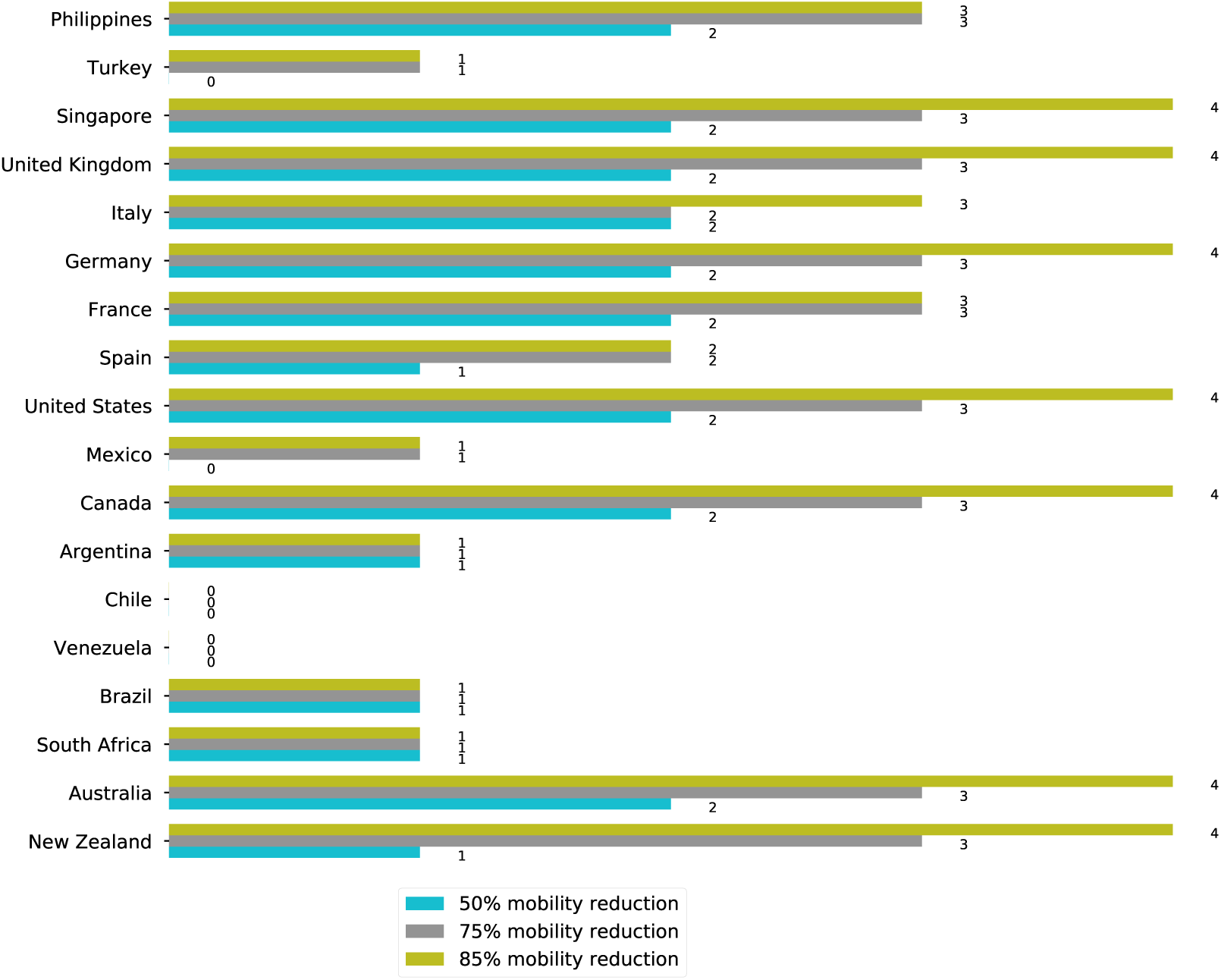
The differences in the predicted onset days between the simulations with reduced mobility flows and the one with regular flow. All simulations are seeded with one case in China on December 1st 2019. Simulated 50%, 75% 85% reductions in mobility flows are applied effectively from December 21st 2019. The predicted onset days are delayed with reduced mobility flows for most countries.

## Discussion

In this work, we augmented a well-established infectious disease model with Google’s aggregate and anonymized mobility data, and demonstrated its application to forecasting the onset of COVID-19 outbreak in a retrospective analysis. Our results suggest that the MA-SEIR model may be used to advise governments or public health organizations for a potential epidemic. It may also play a role in informing a secondary wave of a pandemic, provided corresponding parameters can be estimated. While the dynamics of each patch in the model is based on a simple well-mixed SEIR scheme, the integration of general human movement flows enables infectious disease modeling at a large spatio-temporal scale. This is seen manifested by the global COVID-19 onset forecasting with the first infection case in early December 2019. Additionally, we demonstrated that using the MA-SEIR model to simulate epidemic onset with different levels of mobility reduction offers a framework to understand the relationship between human movement and COVID-19 spread.

It’s worth noting the assumptions in our approach. First, the SEIR model assumes the latent population (“Exposed”) is infectious and that this population will eventually convert to “Infectious” state. Recent studies suggest that for COVID-19 there exists a large undocumented infected population [21] and among them many could be asymptomatic [22]. These findings suggest additional compartments capturing asymptomatic and untested cases may improve the model. Secondly, we assume that the size of each compartmental state in the mobility flow is proportional to the same state in the total population of a patch. This assumption may only hold at the early stage of an epidemic. As quarantines and other measures will likely take place during the progression of the epidemic, the infectious sub-population in the mobility flow will be reduced compared to other sub-populations. Thirdly, our approach requires disease-specific model parameters. In order to provide early warnings, these parameters need to be estimated using early disease transmission information. In our work, the parameters are from [14] where the authors use case numbers in Hubei Province, China from January 16, 2020 to January 25, 2020 to estimate model parameters. Finally, in our retrospective analysis, we use confirmed cases to determine the onset of COVID-19. It’s likely the actual onset is significantly earlier due to undocumented cases and the lag of testing. Therefore, in a real forecasting situation, a further calibration of the predicted onset is required in order to account for these effects.

For our regional simulations, the states with less accurate forecasting results tend to be those that have later epidemic onset. Therefore, one possible approach to improving the forecasting performance is to continuously seed the simulations using confirmed imported cases along with near real-time mobility data as the epidemic progresses. This approach will also likely improve the forecasting performance for New York state, which in our current simulation has no explicit seeding of infection cases. Instead, the infection in New York state is purely seeded by domestic mobility. Recent studies suggest that infection from abroad might have happened in New York state [23]. Therefore explicit seeding of imported cases in New York state is likely to improve its onset prediction.

While the methodology presented in this work is able to forecast epidemic onset with reasonable accuracy, forecasting the magnitude and duration of an epidemic is more challenging and heavily influenced by public health and disease control actions. Further work is required to model the spread of COVID-19 in response to interventions such as social distancing and contact tracing. We speculate that Google’s aggregate and anonymized mobility data can be integrated to more detailed modeling techniques such as stochastic meta-population models and agent-based models, in order to model the COVID-19 pandemic.

## Methods

### Model details

The MA-SEIR metapopulation model is based on earlier SEIR models designed and validated for COVID-19 [13, 14]. In this model we consider each geographic location as a metapopulation. Aggregated anonymized mobility data reported to Google was used to compute the flows between the metapopulations. Unlike classic compartmental models, our approach considers the dynamics of the two following components: (1) the intrinsic dynamics within each metapopulation, and (2) the dynamics due to the flows brought by general human movement.

The SEIR model is often expressed as a continuous-time dynamic system governed by ordinary differential equations (ODEs). Here, we formulate the system as discrete-time Markov process in order to make it compatible with our mobility data, which is indexed by discrete time points:

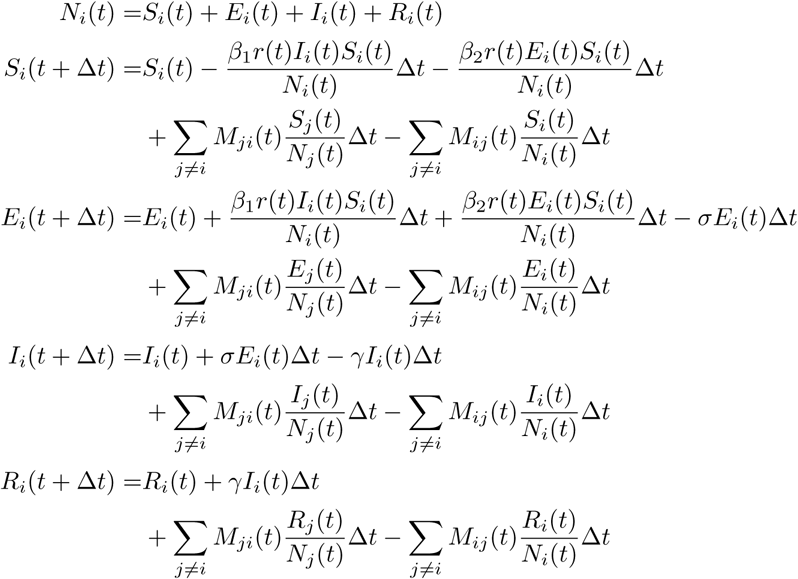

Here, *S*_*i*_(*t*), *E*_*i*_(*t*), *I*_*i*_(*t*) and *R*_*i*_(*t*) are the sizes of the Susceptible, Exposed, Infected and Recovered sub-populations at location *i* at time *t. N*_*i*_(*t*) is the corresponding population. Matrix *M*(*t*) is the mobility matrix, where each entry *M*_*ij*_(*t*) indicates the mobility flow per unit time from location *i* to location *j* at time The parameters *β*_1_ and *β*_2_ are the transmission rates from the Infected and the Exposed, respectively. The parameters *σ* and *γ* are the incubation rate (the reciprocal of incubation period) and the recovery rate (the reciprocal of infection period). Additionally, our model includes a time-dependent multiplier factor *r*(*t*) that is intended for modeling the change of transmission rates over time.

Extensive work has been done in order to estimate the epidemiological parameters for COVID-19 [14, 24–30]. In our retrospective analysis, however we decided to minimize parameter fitting for the following reasons: (1) for a real forecasting task (rather than retrospective analysis) it is unlikely to fit high-quality parameters at the very early stage of an epidemic when reported case numbers are sparse and noisy; and (2) we focused on forecasting the onset of the outbreak, rather than the peak time. Therefore in this work we used a well-established set of parameters that are derived from epidemiological data in Wuhan China in January 2020 [14]. The only fitted parameter in this work is the multiplier factor *r*(*t*), whose value is determined by fitting the early epidemic curves in Italy and South Korea. A single constant *r*(*t*) value is used throughout this work. This is reasonable since at the onset of an epidemiological event, social behavior remains constant until identification. The corresponding basic reproduction number *R*_0_ is 2.99 with corresponding *r*(*t*) value. Table 3 summarizes the parameters of the MA-SEIR model in this work.

**Table 2:** The delay in predicted onset time for global spread.

| **Level of Reductions**<br>**(in Mobility Reduction)** | **Mean of Delay**<br>**(day)** |
| --- | --- |
| 50% | 2.57 |
| 75% | 4.20 |
| 85% | 5.16 |

**Table 3:** The MA-SEIR model parameters. All parameter values except *r*(*t*) are from [14].

| Parameter | Value |
| --- | --- |
| $\beta_1$ (transmission rate from Infected to Susceptible) | $0.15747 \pm 0.01055$ |
| $\beta_2$ (transmission rate from Exposed to Susceptible) | $0.787 \pm 0.053$ |
| incubation period | $7 \pm 2.8$ days |
| infection period | $7 \pm 2.8$ days |
| $\sigma$ (inverse of incubation period) | $0.142857 \pm 0.057143$ |
| $\gamma$ (inverse of infection period) | $0.142857 \pm 0.057143$ |
| $r(t)$ (multiplier factor) | 0.453 |
| $R_0$ (basic reproduction number) | 2.99 |

The ODE solver “odeint” in SciPy [31] was used to solve this system. We derived the initial conditions for each location using mobility data. Seeding was done by further setting Infected size to the number of seeded infections in corresponding locations.

### Mobility data

The Google COVID-19 Aggregated Mobility Research Dataset used for this study is available with permission from Google LLC. This Dataset contains anonymized relative mobility flows aggregated over users who have turned on the Location History setting [19], which is off by default. This is similar to the data used to show how busy certain types of places are in Google Maps — helping identify when a local business tends to be the most crowded. The dataset aggregates flows of people from region to region weekly.

To produce this dataset, machine learning is applied to logs data to automatically segment it into semantic trips [32]. To provide strong privacy guarantees, all trips were anonymized and aggregated using a differentially private mechanism [33, 34] to aggregate flows over time (see [35]). This research is done on the resulting heavily aggregated and differentially private data. No individual user data was ever manually inspected, only heavily aggregated flows of large populations were handled.

All anonymized trips are processed in aggregate to extract their origin and destination location and time. For example, if *n* users traveled from location *a* to location *b* within time interval *t*, the corresponding cell (*a, b, t*) in the tensor would be *n* ± *err*, where *err* is Laplacian noise. The automated Laplace mechanism adds random noise drawn from a zero mean Laplace distribution and yields (*ϵ, δ*)-differential privacy guarantee of *ϵ* = 0.66 and *δ*= 2.1 × 10^*−*29^. The parameter *ϵ* controls the noise intensity in terms of its variance, while *δ* represents the deviation from pure *ϵ*-privacy. The closer they are to zero, the stronger the privacy guarantees.

These results should be interpreted in light of several important limitations. First, the Google mobility data is limited to smartphone users who have opted in to Google’s Location History feature, which is off by default. These data may not be representative of the population as whole, and furthermore their representativeness may vary by location. Importantly, these limited data are only viewed through the lens of differential privacy algorithms, specifically designed to protect user anonymity and obscure fine detail. Moreover, comparisons across rather than within locations are only descriptive since these regions can differ in substantial ways.

In our work and in relation to the dataset described above, the “location” can be a country, state, or county. The “flow” represents the number of users from source to destination locations at the same granularity. The temporal bucket is weekly. Our mobility data doesn’t have sufficient data coverage in China. According to published data, in 2018 Hong Kong has one of the busiest airports in terms of international passenger traffic (74 million) [36], whereas the international passenger traffic volume of Mainland China in 2018 is estimated to be 64 million [37]. We therefore use the mobility data in Hong Kong to approximate the mobility flow originated from China. Regardless of the granularity, all mobility data are protected by the privacy policies described above.

### COVID-19 dataset

Country-level COVID-19 case numbers are obtained from NSSAC (Network Systems Science and Advanced Computing) daily case reports from the University of Virginia [38]. US county-level COVID-19 confirmed case numbers are downloaded from the New York Times [39].

### Experiments

Three experiments are designed to evaluate the performance of the proposed MA-SEIR approach and understand the effects mobility in the spread of COVID-19. We assume that the mobility data beyond the date when the simulation is performed is unknown and can only be estimated.

1. Global country-level modeling of COVID-19 spread is assumed to be performed on January 30 2019 to forecast the epidemic onset, with seeds from reported confirmed cases before this date. The simulation starting date is on December 1st 2019. Therefore the mobility flows used in the simulation includes real-time mobility data for December 2019, January 2020, and approximated mobility for year 2020 (using data from February to May 2019).
2. State-level modeling of COVID-19 spread for United States is assumed to be performed on February 14, 2020, with seeds from reported confirmed cases before this date. The start date of the mobility data is the same as the first date of the simulations. In order to simulate real forecasting scenario, mobility data beyond the date of last seeding infection (February 14, 2020) was replaced by the average mobility data prior to that date.
3. Different levels of simulated reduction of mobility are then incorporated into the MA-SEIR model to evaluate their effects in epidemic onsets. The onset dates derived from these simulations with various degrees of mobility reduction are compared.

An epidemic onset is technically defined as the time when the number of cases is above what is normally expected (“epidemic threshold”) [40]. Different approaches of calculating the epidemic threshold have been proposed [41]. For SARS-CoV-2, there is no consensus epidemic threshold yet. In this work, a subjective fixed epidemic threshold is defined in order to quantify the model performance. We use 100 and 30 cumulative confirmed cases as the epidemic thresholds for countries and states, respectively. To evaluate the model forecasting accuracy, we compute the mean and median of absolute errors in the actual and predicted onset days. We also summarize the positive (early prediction) and negative (late prediction) error individually.

## Data Availability

We do not share any data.

## Acknowledgements

We thank Rif Saurous and Yun Liu for their insightful discussions and advice on this work.

## Supplementary Information

**Table S1:**
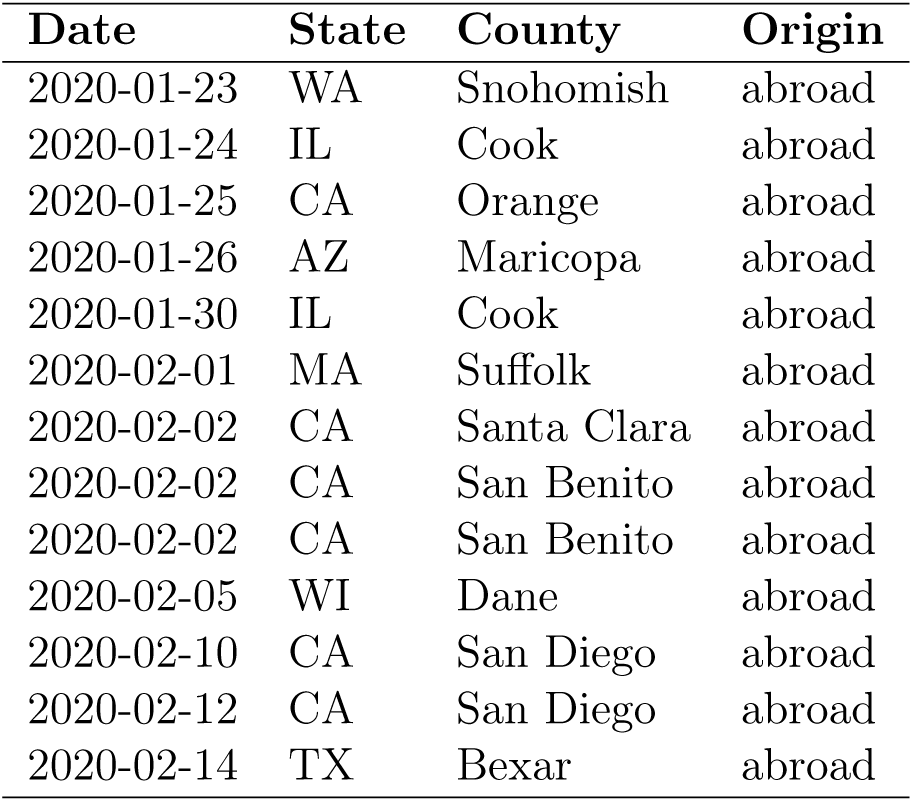
Seeded infection for state-level simulations in the US

| Date | State | County | Origin |
| --- | --- | --- | --- |
| 2020-01-23 | WA | Snohomish | abroad |
| 2020-01-24 | IL | Cook | abroad |
| 2020-01-25 | CA | Orange | abroad |
| 2020-01-26 | AZ | Maricopa | abroad |
| 2020-01-30 | IL | Cook | abroad |
| 2020-02-01 | MA | Suffolk | abroad |
| 2020-02-02 | CA | Santa Clara | abroad |
| 2020-02-02 | CA | San Benito | abroad |
| 2020-02-02 | CA | San Benito | abroad |
| 2020-02-05 | WI | Dane | abroad |
| 2020-02-10 | CA | San Diego | abroad |
| 2020-02-12 | CA | San Diego | abroad |
| 2020-02-14 | TX | Bexar | abroad |

**Table S2:** State-level predicted and reported COVID-19 epidemic onset time in the United States.

| State | Reported Onset | Predicted Onset | Delay (Day) |
| --- | --- | --- | --- |
| Alabama | 2020-03-17 | 2020-04-04 | 18 |
| Alaska | 2020-03-23 | 2020-03-28 | 5 |
| Arizona | 2020-03-19 | 2020-02-19 | -29 |
| Arkansas | 2020-03-18 | 2020-03-28 | 10 |
| California | 2020-03-01 | 2020-02-13 | -17 |
| Colorado | 2020-03-11 | 2020-03-23 | 12 |
| Connecticut | 2020-03-16 | 2020-03-22 | 6 |
| Delaware | 2020-03-20 | 2020-04-20 | 31 |
| Florida | 2020-03-12 | 2020-03-22 | 10 |
| Georgia | 2020-03-11 | 2020-03-25 | 14 |
| Hawaii | 2020-03-20 | 2020-03-25 | 5 |
| Idaho | 2020-03-20 | 2020-03-14 | -6 |
| Illinois | 2020-03-12 | 2020-02-16 | -25 |
| Indiana | 2020-03-18 | 2020-03-21 | 3 |
| Iowa | 2020-03-18 | 2020-03-14 | -4 |
| Kansas | 2020-03-19 | 2020-03-31 | 12 |
| Kentucky | 2020-03-18 | 2020-03-21 | 3 |
| Louisiana | 2020-03-13 | 2020-03-31 | 18 |
| Maine | 2020-03-17 | 2020-03-30 | 13 |
| Maryland | 2020-03-15 | 2020-03-31 | 16 |
| Massachusetts | 2020-03-09 | 2020-02-25 | -13 |
| Michigan | 2020-03-14 | 2020-03-21 | 7 |
| Minnesota | 2020-03-15 | 2020-03-22 | 7 |
| Mississippi | 2020-03-18 | 2020-04-04 | 17 |
| Missouri | 2020-03-19 | 2020-03-13 | -6 |
| Montana | 2020-03-22 | 2020-03-27 | 5 |
| Nebraska | 2020-03-15 | 2020-04-01 | 17 |
| Nevada | 2020-03-16 | 2020-03-16 | 0 |
| New Hampshire | 2020-03-18 | 2020-03-19 | 1 |
| New Jersey | 2020-03-13 | 2020-03-27 | 14 |
| New Mexico | 2020-03-19 | 2020-03-19 | 0 |
| New York | 2020-03-06 | 2020-03-23 | 17 |
| North Carolina | 2020-03-15 | 2020-03-27 | 12 |
| North Dakota | 2020-03-23 | 2020-04-06 | 14 |
| Ohio | 2020-03-15 | 2020-03-24 | 9 |
| Oklahoma | 2020-03-19 | 2020-03-30 | 11 |
| Oregon | 2020-03-14 | 2020-03-18 | 4 |
| Pennsylvania | 2020-03-13 | 2020-03-27 | 14 |
| Rhode Island | 2020-03-18 | 2020-03-19 | 1 |
| South Carolina | 2020-03-16 | 2020-04-04 | 19 |
| South Dakota | 2020-03-25 | 2020-04-11 | 17 |
| Tennessee | 2020-03-14 | 2020-03-26 | 12 |
| Texas | 2020-03-10 | 2020-03-09 | -1 |
| Utah | 2020-03-16 | 2020-03-22 | 6 |
| Vermont | 2020-03-21 | 2020-03-31 | 10 |
| Virginia | 2020-03-14 | 2020-03-27 | 13 |
| Washington | 2020-03-03 | 2020-02-16 | -16 |
| West Virginia | 2020-03-24 | 2020-04-16 | 23 |
| Wisconsin | 2020-03-15 | 2020-02-29 | -15 |
| Wyoming | 2020-03-24 | 2020-04-13 | 20 |

### MA-SEIR parameterization

To evaluate whether the parameters estimated from China can simulate epidemics in other regions, we run simulations about COVID-19 epidemics at fine spatial resolution at US county level. As shown in Figure S1, the resulting curves initially match the confirmed cases. However, as the epidemic develops, the curves start to deviate from the reported case numbers. Two main reasons may contribute to this discrepancy. First, a substantial number of cases were undocumented due to insufficient testing capacity at the beginning of the epidemic [21]. Secondly, control measures implemented at different stages of the epidemic may have led to decreasing transmission rates. We further simulated the epidemics under different scenarios by adjusting the time-dependent multiplier factor *r*(*t*), in order to approximate the decrease of the transmission rates. Figure S1 shows the resulting curves along with confirmed cases in three counties: King County, WA, Santa Clara County, CA, and New York, NY. Indeed, simulations with time-dependent transmission rates produce epidemic curves that more closely match reported positive cases.

Figure S1 shows that time-dependent model parameters are necessary to model the epidemic at longer time scales. However, this is difficult to do early in a new epidemic because it is not clear when control measures will be instituted and what effect they will have on slowing transmission. Additionally, it is complicated by the fact that the number of actual cases is sparse and inconsistently reported, especially early in the epidemic. Nevertheless, we see that a simple MA-SEIR model with time-*independent* parameters can capture the *early* stage of the epidemic reasonably well. When augmented with near real-time mobility data, we hypothesize that the MA-SEIR model can predict the onset of the epidemic in new geographic areas with minimal initial seeding and parameterization. Therefore, this set of parameters are used throughout the simulations at both global country and US state level.

**Figure S1:**
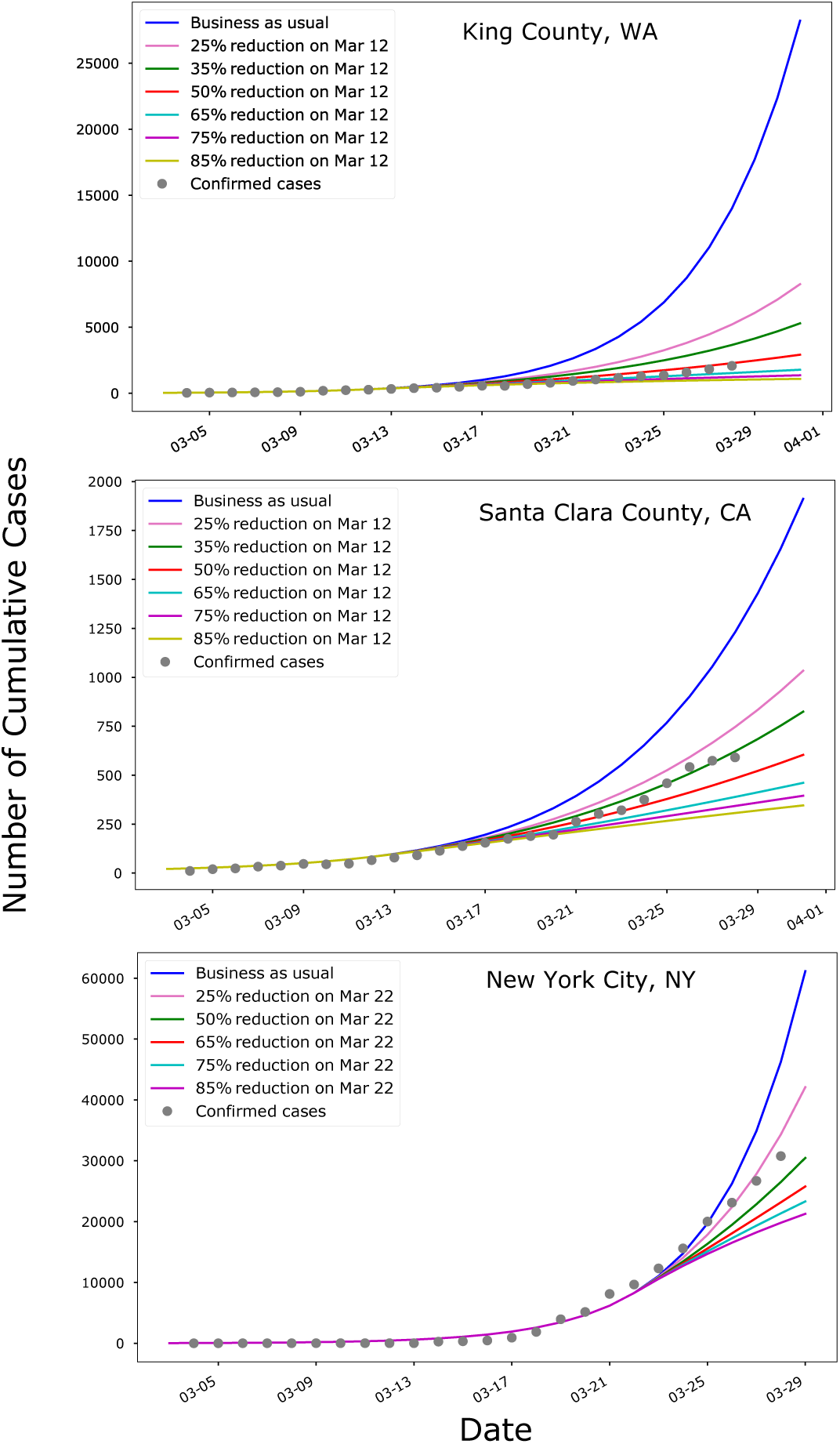
Simulated epidemic curves by MA-SEIR for King County, WA, Santa Clara County, CA and New York City, NY. Each location contains 6 different transmission rate reduction scenarios. The early epidemic curves are well captured by the initial values of the time-dependent parameters in MA-SEIR.

## References

1. World Health Organization COVID-19 Situation Reports https://www.who.int/emergencies/diseases/novel-coronavirus-2019/situation-reports. 2020.

2. Murray, G. D. & Cliff, A. D. A Stochastic Model for Measles Epidemics in a Multi-Region Setting. 2, 158–174. www.jstor.org/stable/621855 (1977).

3. Simini, F., González, M. C., Maritan, A. & Barabási, A.-L. A universal model for mobility and migration patterns. Nature 484, 96–100. issn: 1476-4687 (2012).

4. Colizza, V. & Vespignani, A. Epidemic modeling in metapopulation systems with heterogeneous coupling pattern: Theory and simulations. Journal of Theoretical Biology 251, 450–467. issn: 0022-5193. http://www.sciencedirect.com/science/article/pii/S0022519307005991 (2008).

5. Balcan, D. et al Multiscale mobility networks and the spatial spreading of infectious diseases. Proceedings of the National Academy of Sciences 106, 21484–21489. issn: 0027-8424. eprint: https://www.pnas.org/content/106/51/21484.full.pdf. https://www.pnas.org/content/106/51/21484 (2009).

6. Balcan, D. et al Modeling the spatial spread of infectious diseases: The GLobal Epidemic and Mobility computational model. Journal of Computational Science 1, 132–145. issn: 1877-7503. http://www.sciencedirect.com/science/article/pii/S1877750310000438 (2010).

7. Wesolowski, A. et al Quantifying the Impact of Human Mobility on Malaria. Science 338, 267–270.issn:0036-8075.eprint: https://science.sciencemag.org/content/338/6104/267.full.pdf. https://science.sciencemag.org/content/338/6104/267 (2012).

8. Marshall, J. M. et al Mathematical models of human mobility of relevance to malaria transmission in Africa. Scientific Reports 8, 7713. issn: 2045-2322. https://doi.org/10.1038/s41598-018-26023-1 (2018).

9. Xia, Y., Bjørnstad, O. N. & Grenfell, B. T. Measles Metapopulation Dynamics: A Gravity Model for Epidemiological Coupling and Dynamics. The American Naturalist 164. PMID: 15278849, 267–281. eprint: https://doi.org/10.1086/422341. https://doi.org/10.1086/422341 (2004).

10. Flahault, A. & Valleron, A. A method for assessing the global spread of HIV-1 infection based on air travel. Mathematical Population Studies 3. PMID: 12317173, 161–171. eprint: https://doi.org/10.1080/08898489209525336. https://doi.org/10.1080/08898489209525336 (1992).

11. Hufnagel, L., Brockmann, D. & Geisel, T. Forecast and control of epidemics in a globalized world. Proceedings of the National Academy of Sciences 101, 15124–15129. issn: 0027-8424. eprint: https://www.pnas.org/content/101/42/15124.full.pdf. https://www.pnas.org/content/101/42/15124 (2004).

12. Kraemer, M. U. G. et al The effect of human mobility and control measures on the COVID-19 epidemic in China. Science. issn: 0036-8075. eprint: https://science.sciencemag.org/content/early/2020/03/25/science.abb4218.full.pdf. https://science.sciencemag.org/content/early/2020/03/25/science.abb4218 (2020).

13. Wu, J. T., Leung, K. & Leung, G. M. Nowcasting and forecasting the potential domestic and international spread of the 2019-nCoV outbreak originating in Wuhan, China: a modelling study. The Lancet 395, 689–697. issn: 0140-6736. https://doi.org/10.1016/S0140-6736(20)30260-9 (Feb. 2020).

14. Yang, Z. et al Modified SEIR and AI prediction of the epidemics trend of COVID-19 in China under public health interventions. Journal of Thoracic Disease 12 (2020).

15. Adiga, A. et al Evaluating the impact of international airline suspensions on the early global spread of COVID-19. medRxiv. eprint: https://www.medrxiv.org/content/early/2020/03/02/2020.02.20.20025882.full.pdf (2020).

16. Pei, S. & Shaman, J. Initial Simulation of SARS-CoV2 Spread and Intervention Effects in the Continental US. medRxiv. eprint: https://www.medrxiv.org/content/early/2020/03/27/2020.03.21.20040303.full.pdf. https://www.medrxiv.org/content/early/2020/03/27/2020.03.21.20040303 (2020).

17. Kermack, W. O., McKendrick, A. G. & Walker, G. T. A contribution to the mathematical theory of epidemics. Proceedings of the Royal Society of London. Series A, Containing Papers of a Mathematical and Physical Character 115, 700–721 (1927).

18. Hethcote, H. W. The Mathematics of Infectious Diseases. SIAM Review 42, 599–653 (Jan. 2000).

19. Google Account Help: Manage Your Location History https://support.google.com/accounts/answer/3118687. 2020.

20. Huang, C. et al Clinical features of patients infected with 2019 novel coronavirus in Wuhan, China. The Lancet 395, 497–506 (2020).

21. Li, R. et al Substantial undocumented infection facilitates the rapid dissemination of novel coronavirus (SARS-CoV2).

22. Mizumoto, K., Kagaya, K., Zarebski, A. & Chowell, G. Estimating the asymptomatic proportion of coronavirus disease 2019 (COVID-19) cases on board the Diamond Princess cruise ship, Yokohama, Japan, 2020. Eurosurveillance 25. https://www.eurosurveillance.org/content/10.2807/1560-7917.ES.2020.25.10.2000180 (2020).

23. Gonzalez-Reiche, A. S. et al Introductions and early spread of SARS-CoV-2 in the New York City area. medRxiv. eprint: https://www.medrxiv.org/content/early/2020/04/16/2020.04.08.20056929.full.pdf.https://www.medrxiv.org/content/early/2020/04/16/2020.04.08.20056929 (2020).

24. Li, Q. et al Early Transmission Dynamics in Wuhan, China, of Novel Coronavirus–Infected Pneumonia. New England Journal of Medicine 382. PMID: 31995857, 1199–1207. eprint: https://doi.org/10.1056/NEJMoa2001316. (2020).

25. Liu, Y., Gayle, A. A., Wilder-Smith, A. & Rocklöv, J. The reproductive number of COVID-19 is higher compared to SARS coronavirus. Journal of Travel Medicine 27. taaa021. issn: 1195-1982. eprint: https://academic.oup.com/jtm/article-pdf/27/2/taaa021/32902430/taaa021.pdf. https://doi.org/10.1093/jtm/taaa021 (Feb. 2020).

26. Liu, T. et al. Time-varying transmission dynamics of Novel Coronavirus Pneumonia in China. bioRxiv. eprint: https://www.biorxiv.org/content/early/2020/02/13/2020.01.25.919787.full.pdf. https://www.biorxiv.org/content/early/2020/02/13/2020.01.25.919787 (2020).

27. Li, J. et al Estimation of the epidemic properties of the 2019 novel coronavirus: A mathematical modeling study. medRxiv. eprint: https://www.medrxiv.org/content/early/2020/02/20/2020.02.18.20024315.full.pdf. https://www.medrxiv.org/content/early/2020/02/20/2020.02.18.20024315 (2020).

28. Wang, M. & Qi, J. A deterministic epidemic model for the emergence of COVID-19 in China. medRxiv. eprint: https://www.medrxiv.org/content/early/2020/03/10/2020.03.08.20032854.full.pdf. https://www.medrxiv.org/content/early/2020/03/10/2020.03.08.20032854 (2020).

29. Kucharski, A. J. et al Early dynamics of transmission and control of COVID-19: a mathematical modelling study. The Lancet Infectious Diseases. issn: 1473-3099. https://doi.org/10.1016/S1473-3099(20) 30144-4 (XXXX).

30. Lai, C.-C., Shih, T.-P., Ko, W.-C., Tang, H.-J. & Hsueh, P.-R. Severe acute respiratory syndrome coronavirus 2 (SARS-CoV-2) and coronavirus disease-2019 (COVID-19): The epidemic and the challenges. International Journal of Antimicrobial Agents 55, 105924. issn: 0924-8579. http://www.sciencedirect.com/science/article/pii/S0924857920300674 (2020).

31. Virtanen, P. et al SciPy 1.0: Fundamental Algorithms for Scientific Computing in Python. Nature Methods 17, 261–272 (2020).

32. Kirmse, A., Udeshi, T., Bellver, P. & Shuma, J. Extracting patterns from location history in Proceedings of the 19th ACM SIGSPATIAL International Conference on Advances in Geographic Information Systems (2011), 397–400.

33. Wilson, R. et al Differentially Private SQL with Bounded User Contribution in (2020).

34. Dwork, C., Roth, A., et al The algorithmic foundations of differential privacy. Foundations and Trends® in Theoretical Computer Science 9, 211–407 (2014).

35. HOW GOOGLE ANONYMIZES DATA https://policies.google.com/technologies/anonymization.

36. ACI World releases preliminary 2018 world airport traffic rankings Passenger traffic: Passenger traffic remains resilient but cargo hubs see volume growth weaken India becomes world’s third largest aviation market for passenger traffic. https://aci.aero/news/2019/03/13/preliminary-world-airport-traffic-rankings-released/. 2019.

37. Statistics of Key Performance Indicators for China’s Civil Aviation Industry in December 2018 http://www.caac.gov.cn/en/HYYJ/SJ/201902/W020190226511539703856.pdf. 2019.

38. COVID-19 Surveillance Dashboard Biocomplexity Institute, University of Virginia https://nssac.bii.virginia.edu/covid-19/dashboard/. 2020.

39. The New York Times COVID-19 Trancking Page https://www.nytimes.com/interactive/2020/us/coronavirus-us-cases.html. 2020.

40. Principles of epidemiology in public health practice: an introduction to applied epidemiology and biostatistics https://www.cdc.gov/csels/dsepd/ss1978/index.html. 2012.

41. Cai, J. et al A maximum curvature method for estimating epidemic onset of seasonal influenza in Japan. BMC Infectious Diseases 19, 181. issn: 1471-2334. https://doi.org/10.1186/s12879-019-3777-x (Feb. 2019).

